# Projected Health and Economic Effects of Expanded Access to Anti-Obesity Medications in Medicare

**DOI:** 10.64898/2026.10.26.26366542

**Authors:** Alison Sexton Ward, PhuongGiang Nguyen, Jack Chapel, Bryan Tysinger, Erin Trish, Darius Lakdawalla, Dana Goldman

## Abstract

**Background:** The CMS GLP-1 Bridge demonstration (July 2026–December 2027) expands access to GLP-1s for additional eligible Medicare Part D beneficiaries for a $50 monthly copay. Its long-run value depends on uptake, discontinuation, and which beneficiaries initiate therapy, in addition to policy decisions.

**Methods:** We used the Future Elderly Model, a microsimulation of Americans aged 55 and older, to model the costs and benefits of expanded GLP-1 access to the newly Bridge-eligible population, assuming that it continues over a 10-year horizon and is accompanied by reduced GLP-1 pricing taking effect for already covered indications in Part D beginning in 2028. We compared a status quo with two expanded-access scenarios: high discontinuation paired with high reinitiation (Rodriguez et al.), and low discontinuation paired with low reinitiation (Understanding America Study). Each scenario was run for two different definitions of the Bridge-eligible population – a broad definition that includes beneficiaries with prior myocardial infarction or stroke and a narrow definition that excludes them.

**Results:** Expanded access generated positive net social value in every scenario, from $44.3 billion to $58.5 billion over 10 years. Medicare net spending fell in every scenario, with cumulative savings of $21.2–27.1 billion, including $31.9 billion in diabetes drug price savings. Return on investment was 117–216% for Medicare and 198–346% for society. The eligible population mattered more than persistence. Net social value was 23–29% higher in the narrow population, because its initiators are healthier and have more to gain form access to GLP-1 treatment. Low discontinuation raised net social value by only 2–7% but raised Medicare drug spending, so ROI was higher under high discontinuation.

**Conclusions:** Expanded Medicare access to GLP-1s is projected to produce substantial social value across a range of persistence and eligibility assumptions. Extending the lower demonstration price to previously covered indications would allow the program to pay for itself from Medicare’s perspective. Because health gains accrue over many years, evaluating the demonstration only over its 18-month window would understate its benefits.

## Introduction

Glucagon-like peptide-1 (GLP-1) receptor agonists have transformed the treatment of obesity. In clinical trials, these therapies have produced sustained weight loss of 15–20% or more, along with improvements in cardiovascular risk factors, glycemic control, and other obesity-related conditions.^1-7^ Access, however, has not kept pace with the accumulating evidence on impact.

Coverage of GLP-1s prescribed for weight loss remains limited in both public and private insurance.^8^ Many payers have been reluctant to expand coverage because of cost and uncertainty about whether patients will stay on treatment (or with the insurer) long enough for the benefits to offset it.^9^ Fewer than one in five employers cover GLP-1s for obesity.^8^

Medicare has historically been prohibited by statute from covering medications used for weight loss, despite the substantial health and economic benefits that could come from doing so.^4^ In December 2025, the Centers for Medicare and Medicaid Services (CMS) announced the Better Approaches to Lifestyle and Nutrition for Comprehensive hEalth (BALANCE) Model, a voluntary Medicare and Medicaid model to test whether expanding access to GLP-1s and evidence-based lifestyle supports for weight loss could improve health outcomes without generating unsustainable costs.^10^ In Medicare, CMS initially proposed a two-part approach to expanded GLP-1 access, including a six-month “Bridge” beginning in July 2026 that would operate outside the Medicare Part D program, followed by a model within the Part D program that would run from January 2027 through December 2031 conditional on participation from manufacturers and Part D plans. Manufacturers agreed to provide GLP-1s at a monthly net price of $245 (including beneficiary cost-sharing) for both the expansion population newly eligible for GLP-1 coverage and for beneficiaries being treated by indications already covered under Part D (e.g., diabetes). However, plan participation did not meet the minimum required threshold of 80% of total national Part D enrollment, due to uncertainty around utilization, selection, and risk protection.^11^

In April 2026, CMS announced the indefinite delay of the Medicare BALANCE model and extended Bridge through the end of 2027. The Bridge demonstration operates outside the standard Part D benefit, instead using a central processor to manage prior authorization, claims, and pharmacy payment. Bridge extends GLP-1 coverage beyond Medicare beneficiaries with diabetes and other previously covered conditions, opening eligibility to three additional groups: (1) beneficiaries with a body mass index (BMI) of at least 35; (2) beneficiaries with a BMI of at least 30 and a diagnosis of heart failure, uncontrolled hypertension, or chronic kidney disease stage 3a or above; and (3) beneficiaries with a BMI of at least 27 and a diagnosis of prediabetes, previous myocardial infarction, previous stroke, or symptomatic peripheral artery disease.^12^ According to KFF estimates, this expands coverage to roughly 3.8 million newly-eligible Medicare beneficiaries, with anticipated benefits including improved health outcomes, reduced incidence of chronic disease, and lower downstream health care utilization.^13,14^ Beneficiaries pay a $50 monthly copayment.^15^

CMS noted that the extended Bridge timeline “would offer a valuable opportunity to provide plans with additional data and facilitate a smoother transition to the potential implementation of BALANCE in Part D.”^16^ In this paper, we model the costs and benefits of expanded GLP-1 access to the three newly eligible groups describe above, assuming that it continues over a 10-year horizon and is accompanied by reduced GLP-1 pricing for already covered indications beginning in 2028.

We build on prior work by modeling a range of discontinuation and reinitiation scenarios. Claims-based studies report wide variation in first-year discontinuation, ranging from 24% to 65% depending on the population, therapy, and definition used.^17,18^ However, these estimates are drawn from claims-based studies of insured populations and do not capture cash-pay or out-of-network use, leaving open the question of how representative they are of real-world persistence, particularly among the uninsured and underinsured patients who stand to be most affected by coverage expansions.

We use new survey evidence on discontinuation from the nationally representative Understanding America Study (UAS) that addresses some of the limitations in prior claims-based studies.^19^ Using data that capture both insured and cash-pay GLP-1 use, the survey finds lower discontinuation rates than most claims-based estimates—12.8% by 6 months, 22.1% by 12 months, and 33.4% by 24 months—and identifies out-of-pocket cost, rather than side effects, as one of the strongest predictors of discontinuation, on par with vomiting and larger than other commonly cited tolerability concerns. Notably, patients using GLP-1s primarily for weight loss were more likely to discontinue than those using them to treat diabetes, consistent with the former group facing less consistent coverage. Taken together, this evidence suggests that observed adherence patterns may reflect financial access as much as—or more than—clinical tolerability, and that expanding coverage would likely improve persistence.

However, the magnitude and timing of these benefits—and how they weigh against program costs—depend critically on assumptions about uptake and adherence that remain uncertain. We use the Future Elderly Model (FEM), a microsimulation of the health and health care spending of older Americans, to project outcomes for Medicare beneficiaries under a status quo scenario and two “Bridge to BALANCE” demonstration treatment scenarios that differ in GLP-1 discontinuation. For each scenario, the model tracks GLP-1 initiation, discontinuation, reinitiation, and weight regain, and translates changes in BMI into changes in chronic disease, quality of life, and health care spending. By varying adherence assumptions systematically, we characterize how sensitive the program’s projected value is to the adherence concerns that have historically limited GLP-1 coverage. Our analysis provides a framework that payers and policymakers can use to evaluate similar coverage expansions going forward.

## Methods

The FEM is an economic-demographic microsimulation of the health and economic outcomes of older Americans, developed with funding from the Centers for Medicare & Medicaid Services, the National Institute on Aging, the Department of Labor, and the MacArthur Foundation. FEM draws on three nationally representative surveys: the Health and Retirement Study (HRS), the Medical Expenditure Panel Survey (MEPS), and the Medicare Current Beneficiary Survey (MCBS). Combining these sources provides more comprehensive information for the model’s development than any single survey can provide. FEM advances in two-year cycles that mirror the frequency of the HRS. In each cycle, it simulates individuals’ health outcomes, total health care spending, and GLP-1 treatment costs (Figure 1). A full technical description is available online.^20^

**Figure 1.**
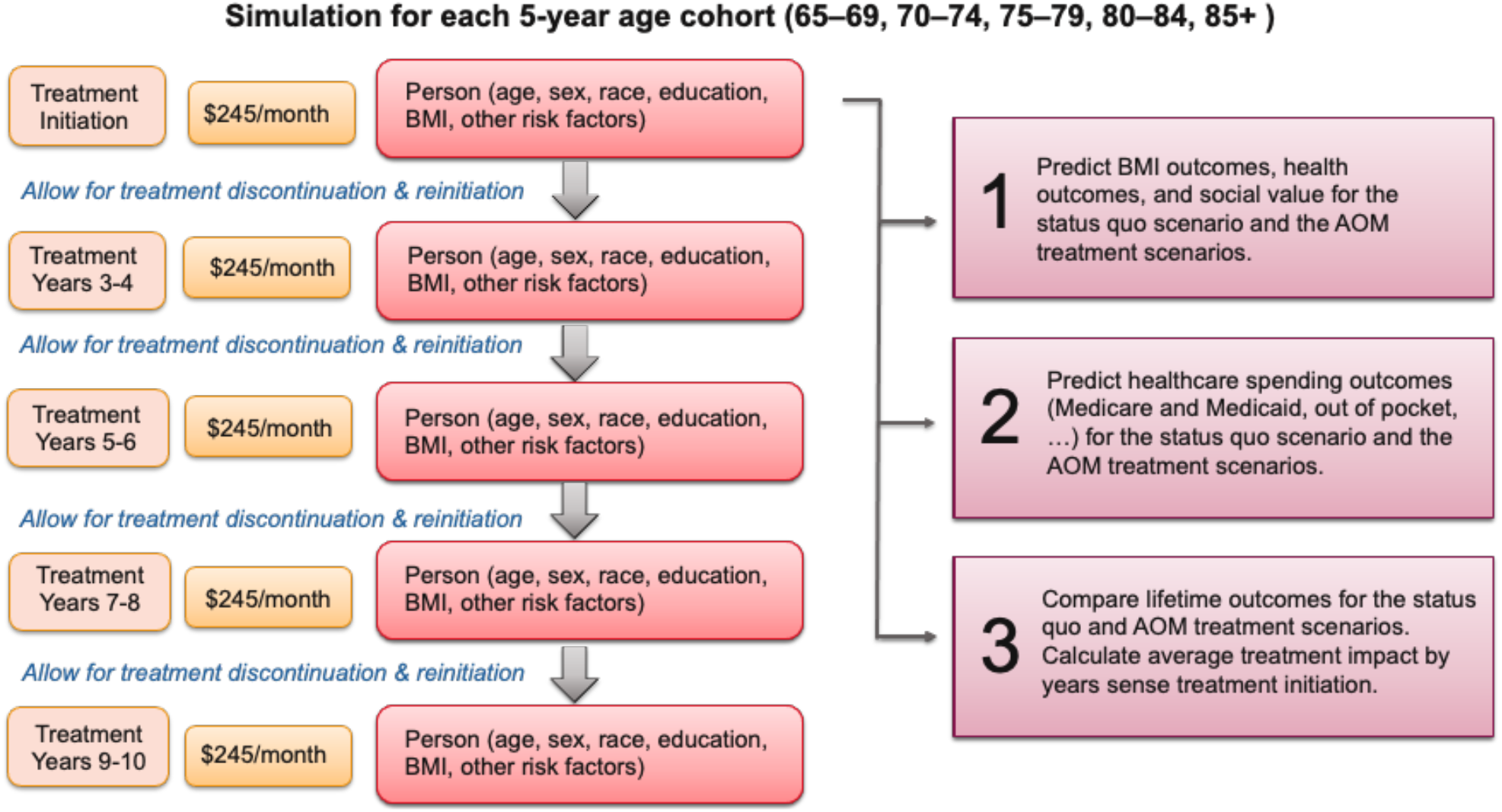
Structure of the microsimulation model.

Health transitions are estimated as first-order Markov processes that depend on age, sex, education, race/ethnicity, existing health conditions, BMI, and functional status. Chronic conditions—heart disease, stroke, diabetes, cancer, and lung disease—are self-reported in the HRS and treated as absorbing states: once a simulated individual develops a condition, they retain it until death. BMI is modeled as a function of age, sex, education, race/ethnicity, marital status, health conditions, functional limitations, smoking status, and prior BMI.

Predicted health status feeds into estimates of health care spending, which is modeled as a function of age, sex, race/ethnicity, education, and chronic conditions. For Medicare beneficiaries age 65 and older, estimates are based on the MCBS and additionally include functional limitations, nursing home residency, and mortality. These estimates capture all health care expenditures except GLP-1 treatment costs, which we model separately.

### Study population and “Bridge to BALANCE” Demonstration eligibility

We simulated a nationally representative cohort of Americans aged 65 and older. Using FDA-labeled indications, we first identified all individuals clinically eligible for GLP-1 treatment for weight management. We then separated them into beneficiaries who already have GLP-1 coverage through Medicare Part D, because of type 2 diabetes or moderate-to-severe obstructive sleep apnea^a^, and beneficiaries without such coverage. Among the latter, we identified those who gain access through the Medicare GLP-1 Bridge demonstration (hereafter, Bridge) by applying CMS’s clinical criteria.^12,b^

Established cardiovascular disease creates an overlap between Part D and Bridge eligibility. Wegovy is covered under standard Part D to reduce major adverse cardiovascular events in patients with established cardiovascular disease and overweight or obesity, while the Bridge criteria list previous myocardial infarction, previous stroke, and symptomatic peripheral artery disease as qualifying diagnoses for beneficiaries with a BMI of at least 27.^12^ CMS acknowledges this overlap. Because the pathway a patient uses depends on how the prescription is documented, we include these beneficiaries as Bridge-eligible in our main analysis labeled “broad” to reflect the added beneficiaries, and test their exclusion in a sensitivity analysis labeled “narrow.” Specifically, in the narrow scenario, beneficiaries with prior myocardial infarction or stroke are excluded from the eligible Bridge population, but initiation estimates are unchanged, so the number of eligible beneficiaries starting GLP-1 is the same and only their health profile differs.

### Simulation design and scenarios

The simulation begins in 2027 and projects health, health care spending, treatment costs, and other economic outcomes over a 10-year horizon, allowing us to compare results across alternative treatment scenarios.^c^ For each Bridge-eligible beneficiary, we simulate outcomes with and without Bridge access, assuming the expanded access does not expire in December 2027 and, rather, continues for the full 10-year model horizon. We consider three scenarios: a status quo scenario and two Bridge expanded-access scenarios that differ in GLP-1 discontinuation rates (high and low).

#### Status quo

This scenario assumes the Bridge demonstration does not exist and that Medicare continues to exclude coverage of GLP-1s for weight management for the beneficiaries modeled in our simulation. Historical GLP-1 use rates are held constant over the 10-year horizon.

#### Expanded access scenarios

In the two expanded-access scenarios, beneficiaries gain coverage for a GLP-1 therapy that reduces BMI, reflecting mean weight loss in recent clinical trials of higher-dose regimens.^6,7^ After the initial reduction, FEM’s empirically based BMI transition model predicts participants’ weight in subsequent cycles while on GLP-1 treatment. Treatment initiation is limited to those younger than age 85. We also model a reduction in diabetes incidence consistent with clinical trial results.^21,22^ GLP-1 therapy reduces diabetes risk indirectly via BMI, as well as directly. The total (direct and indirect) diabetes effect was calibrated to match the reduction in diabetes risk from the clinical trial for the first two model cycles (four years) while on treatment, approximating the three-year time horizon of the trial. After two model cycles, the natural diabetes transition resumes. We do not extrapolate the direct diabetes effect beyond four years, as limited evidence exists on long-term effect of GLP-1 therapy on glycemic status and diabetes risk. Treatment discontinuation at any point during a model cycle completely eliminates the direct diabetes risk reduction.

Long-term persistence remains a central uncertainty in modeling GLP-1 use, thus we incorporated literature-based estimates of discontinuation and reinitiation in our two Bridge scenarios. We model quarterly discontinuation rates as functions of time on treatment and time since discontinuation, using probabilities from two sources. The goal is to represent the range of discontinuation estimates from the literature, and each scenario is run with both the broad and the narrow patient populations.

##### High discontinuation & high reinitiation

Rodriguez et al. (2025) is a retrospective cohort study of U.S. adults with overweight or obesity who initiated GLP-1 therapy between 2018-2023.^23^ The study found lower discontinuation among users with diabetes (46.5% within one year) than among users with obesity (64.8%). Differences in coverage and out-of-pocket costs may drive this gap, and Bridge addresses both directly. Thus, our high discontinuation scenario uses the Rodriguez et al. rates among patients with type 2 diabetes – 47% discontinued within one year and 64% within two years, and 47% of those who discontinued reinitiated within one year post discontinuation and 57% within two years. (The literature-based discontinuation parameters are reported in Appendix Table A1.)

##### Low discontinuation & low reinitiation

Claims-based studies miss medications purchased out of pocket or obtained outside traditional insurance channels. This is an important limitation because coverage for obesity remains highly variable and fewer than one in five employers cover GLP-1s prescribed primarily for obesity.^8^ Our low discontinuation scenario therefore uses data from the Understanding America Study (UAS), a nationally representative internet panel of U.S. adults, in which 33% of respondents using GLP-1 medications discontinued within two years and 30% of users reported treatment gaps longer than 3 months which we associate with reinitiation.^19^ The data are self-reported rates of discontinuation and therefore they reflect a broader definition of persistence which may include beneficiaries who take short gaps in treatment.

The two sources differ in data type and in how discontinuation is defined, claims-based versus self-reported, so together they bracket a reasonable range of real-world GLP-1 use. Because stopping and restarting treatment are linked behaviors, each scenario pairs a discontinuation rate with the reinitiation rate observed in the same source. Differences between the scenarios therefore reflect the combined effect of persistence and cycling on and off treatment, not discontinuation alone.

##### BMI trajectories

GLP-1 therapy results in weight loss following an exponential decay trajectory, with weight loss plateauing at approximately 20% from baseline by two years after treatment initiation, approximating recent clinical trials.^6,22^ After GLP-1 discontinuation, simulated individuals’ weight returns toward its pre-treatment level along a trajectory estimated in a 2026 systematic review and nonlinear meta-regression of published trial data.^24^ The fitted exponential recovery curve shows that weight consistently climbs back after treatment cessation, with 60% of the weight lost during treatment recovered after one year and a plateau at 75.3% of the loss by the second year. We applied these curves to calculate quarterly weight changes.

### GLP-1 uptake rate

We modeled GLP-1 initiation among the newly eligible Medicare population, allowing rates to vary by 5-year age group (65–69, 70–74, 75–79, and 80–84) and by simulation year. Our main-analysis rates draw on early reports of Bridge enrollment^25^ and on treatment interest expressed in the UAS. The simulation begins with no GLP-1 users, so first-year initiation must build the initial stock of users. We set 2027 prevalence of GLP-1 use to match UAS rates of current and former use among similarly aged respondents without diabetes: 8% for ages 65–69, 7% for ages 70–74, 5% for ages 75–79, and 4% for ages 80–84. Reaching these levels requires first-time initiation among 9.0%, 7.8%, 5.6%, and 4.5% of each age group, respectively to account for discontinuation. Prevalence then rises over the next three years to a ceiling equal to current and former use plus the share of non-users who report interest in GLP-1 therapy: 16%, 13%, 9%, and 7%, respectively. We hold prevalence at this ceiling through 2036. Initiation rates are calibrated to reach and then sustain these prevalence targets given the persistence and reinitiation assumptions described below. We do not model treatment initiation at age 85 and older, consistent with the low use rates reported in the UAS for this age group.

### GLP-1 treatment costs and price extension to previously covered indications

Participating manufacturers agreed to provide GLP-1 therapies through Bridge at a net price of $245 per monthly supply. Medicare pays $195 per month, and beneficiaries pay a $50 monthly copay.^15^ We assume these prices and copays would continue for the full 10 years. The current 18-month demonstration applies this price only to prescriptions for weight management and does not extend it to Part D-covered indications. However, BALANCE would extend this pricing to GLP-1s prescribed for those indications (type 2 diabetes, MASH, and obstructive sleep apnea) covered under Part D.^26^ In our model, we also extend these pricing reductions to previously covered indications beginning in 2028 (model year 2), which generates Medicare savings on existing prescriptions. We estimated these savings by multiplying the number of beneficiaries who access GLP-1s through existing Part D coverage by the difference between current net prices and the Bridge price. Current net prices are the CMS-negotiated maximum fair price (MFP) for Ozempic (slated to take effect in 2027) and the current net price for Mounjaro.^27^ We used Medicare population estimates from the Trustees’ Report and assumed constant diabetes prevalence over the 10-year horizon.^28^ Based on ASPE estimates of GLP-1 use among beneficiaries with diabetes^15^, we projected that use grows from 15% to 25% over the simulation horizon.

### Simulation Outcomes

We simulate each 5-year age cohort under all three scenarios and report cumulative differences between each Bridge scenario (high and low discontinuation) and the status quo for the Medicare population. Outcomes include Medicare spending, Medicare GLP-1 costs, beneficiary out-of-pocket GLP-1 costs, life years lived with diabetes, and net social value. To capture quality-of-life gains from expanded access to GLP-1s, we use Generalized-Risk-Adjusted Cost-Effectiveness (GRACE), which, unlike traditional cost-effectiveness analysis, complies with the Inflation Reduction Act’s prohibition on discriminatory value assessment methods.^29,30^ Net social value equals (1) the value of gains in longevity and quality of life, plus (2) lifetime disability expenditure savings, minus (3) incremental lifetime net health care spending, defined as treatment costs minus health care cost offsets. We value each year in perfect health at $150,000, a conventional assumption,^31^ and discount all costs and benefits at 3% annually, except in internal rate of return (IRR) calculations, which require no discount rate.^32^ We do not model Part D plan behavior.

## Results

Table 1 reports 10-year cumulative outcomes for all four scenarios, and Figures 2–4 show how those outcomes evolve over time. In 2027, UAS-based uptake rates generated almost 740,000 first-time GLP-1 initiators, growing to a cumulative 2.35 million by 2036. Many of these beneficiaries go on to discontinue, and some reinitiate.

**Table 1.** Ten-year outcomes of the expanded GLP-1 access.

| Outcome (2027–2036,<br>cumulative) | Broad eligibility |  | Narrow eligibility |  |
| --- | --- | --- | --- | --- |
|  | Low<br>discontinuation<br>/ low<br>reinitiation | High<br>discontinuation<br>/ high<br>reinitiation | Low<br>discontinuation<br>/ low<br>reinitiation | High<br>discontinuation<br>/ high<br>reinitiation |
| **Treatment and health outcomes** |  |  |  |  |
| Cumulative first-time initiators<br>(millions) | 2.35 | 2.35 | 2.35 | 2.35 |
| Years lived with diabetes avoided<br>(thousands) | 1,087 | 753 | 1,242 | 860 |
| **Spending outcomes** |  |  |  |  |
| Medicare GLP-1 drug spending | 18.1 | 12.8 | 18.5 | 12.5 |
| Medical cost offsets | 7.4 | 5.7 | 10.0 | 7.8 |
| Diabetes drug price savings | 31.9 | 31.9 | 31.9 | 31.9 |
| Net CMS spending | (21.2) | (24.8) | (23.4) | (27.1) |
| Net social value | 45.2 | 44.3 | 58.5 | 54.6 |

**Notes:** Results are for Bridge-eligible beneficiaries and include diabetes drug price savings beginning in 2028. Broad eligibility includes beneficiaries with prior myocardial infarction or stroke; narrow eligibility excludes them. Low discontinuation is paired with low reinitiation and high discontinuation with high reinitiation. Values are cumulative over 2027–2036 [discounted at 3%]. Net CMS spending equals Medicare drug spending minus medical cost offsets and minus diabetes drug price savings. Parentheses denote negative values; negative net CMS spending indicates net savings to Medicare. Components may not sum to totals because of rounding.

**Figure 2.**
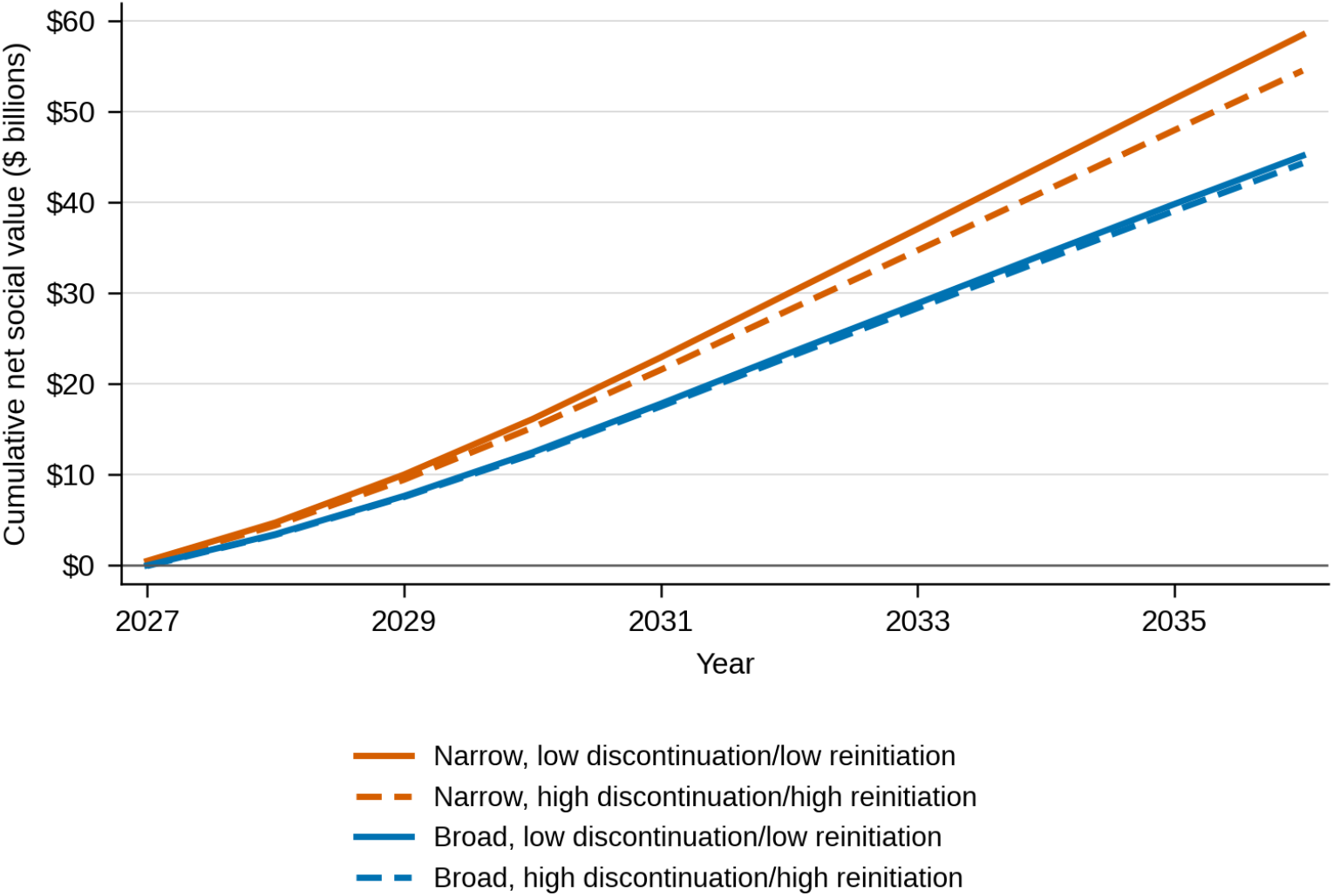
Cumulative net social value, 2027–2036. **Notes:** Results are for Bridge-eligible beneficiaries and include diabetes drug price savings beginning in 2028. Scenarios combine eligibility (broad or narrow) with discontinuation and reinitiation (low discontinuation paired with low reinitiation, or high with high).

### Net Social Value

Across all discontinuation/reinitiation scenarios (high discontinuation paired with high reinitiation and low discontinuation paired with low reinitiation) and eligible populations (the broad population includes beneficiaries with prior myocardial infarction or stroke; the narrow population excludes them), the Bridge demonstration generated positive cumulative net social value over the 10-year horizon (Figure 2). Net social value was positive in every year of every scenario except 2027 in the high discontinuation scenario with the broad population, where it was slightly negative (−$42 million) before turning positive in 2028.

Cumulative net social value by 2036 ranged from $44.3 billion (high discontinuation, broad) to $58.5 billion (low discontinuation, narrow) (Table 1; Figure 2). The population definition mattered more than treatment persistence. Net social value under the narrow definition was 23– 29% higher than that under the broad definition ($54.6–58.5 billion versus $44.3–45.2 billion). Because the number of initiators is the same in both populations, this difference reflects who initiates. The broad population includes beneficiaries with prior myocardial infarction or stroke, who have already developed conditions that GLP-1s might otherwise prevent and who face higher mortality, so each initiator has less to gain. In the narrow population, initiators are healthier on average, and GLP-1s have more potential to prevent chronic conditions and other obesity-related comorbidities.

In contrast, low discontinuation raised net social value by only 2% in the broad population ($45.2 billion versus $44.3 billion) and 7% in the narrow population ($58.5 billion versus $54.6 billion). Diabetes drug price savings, which total $31.9 billion by 2036 and are identical across scenarios, do not contribute to the differences between scenarios. Because each discontinuation scenario is paired with the reinitiation rate observed in the same source, high reinitiation in the high discontinuation scenario partly offsets the treatment lost to discontinuation, and low reinitiation in the low discontinuation scenario partly offsets the benefit of staying on treatment. This pairing may help explain why persistence had a modest effect on net social value, and these differences reflect the combined effect of persistence and cycling on and off treatment rather than discontinuation alone.

### Medicare budget impact

We report the budget impact of the Demonstration assuming reduced pricing applies to GLP-1s prescribed for weight management under the expanded eligibility criteria (beginning in 2027) and also applies to GLP-1s for diabetes and other previously covered indications beginning in 2028, generating savings on existing prescriptions.

The lower GLP-1 prices would generate net savings to Medicare in every scenario (Figure 3). Cumulative net spending turned negative (such that Medicare was saving money) in 2028 under high discontinuation/high reinitiation and in 2029 under low discontinuation/low reinitiation. By 2036, cumulative net savings reached $21.2–27.1 billion. The diabetes price savings ($31.9 billion by 2036) were the same in all scenarios. Savings were larger under high discontinuation/high reinitiation (for example, $24.8 billion versus $21.2 billion in the broad population) because Bridge drug spending was lower.

**Figure 3.**
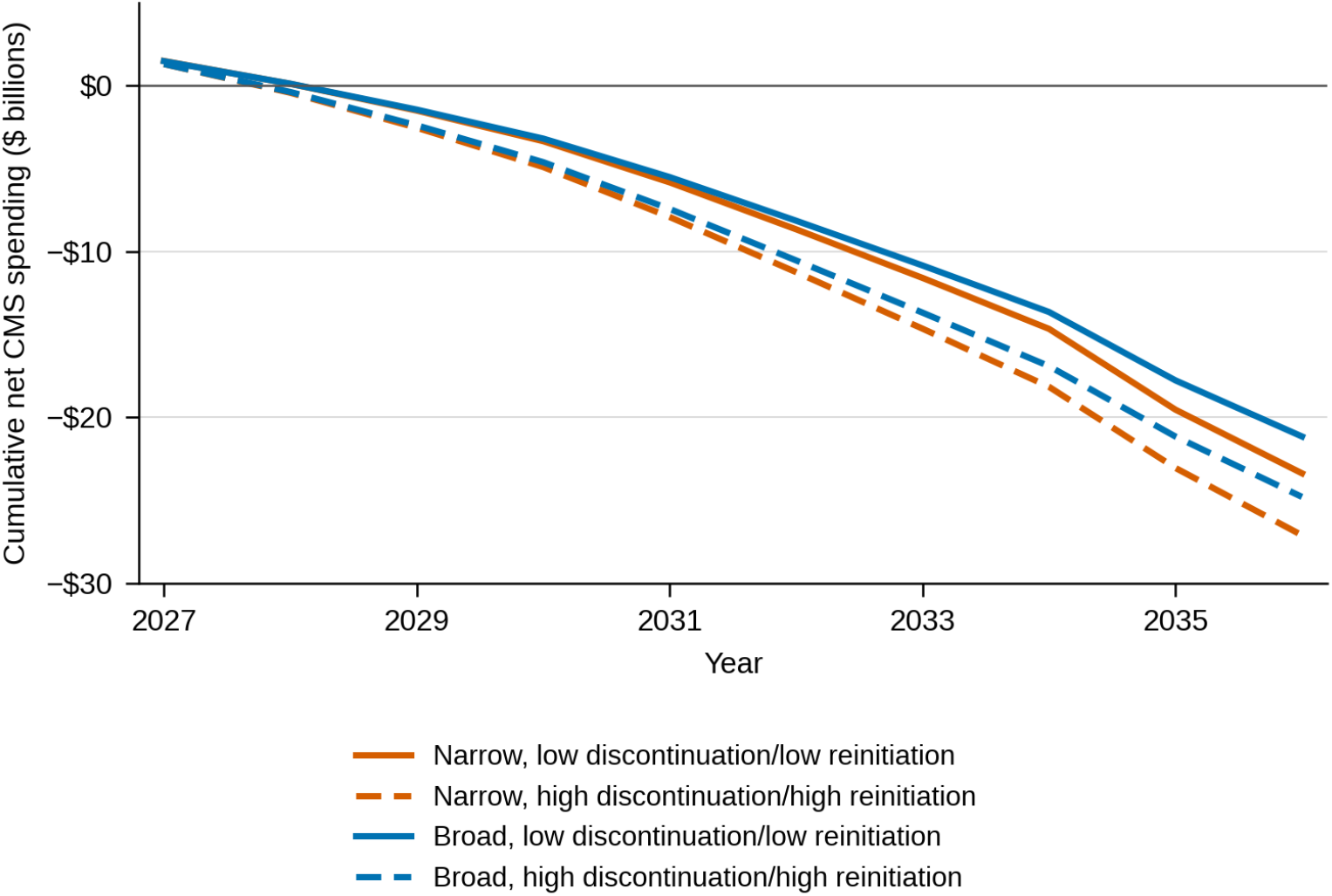
Cumulative net CMS spending, 2027–2036. **Notes:**Results are for Bridge-eligible beneficiaries and include diabetes drug price savings beginning in 2028. Scenarios combine eligibility (broad or narrow) with discontinuation and reinitiation (low discontinuation paired with low reinitiation, or high with high). Positive values indicate added spending and negative values indicate savings.

### Return on investment

We calculated 10-year cumulative return on investment (ROI) from two perspectives (Figure 4). The CMS ROI measures Medicare’s return on its Bridge GLP-1 spending. It is the reduction in other Medicare spending (medical cost offsets) plus diabetes drug price savings, net of Bridge drug spending, relative to Bridge drug spending. The social ROI measures the return to society on GLP-1 treatment spending: net social value per dollar of total spending on GLP-1 treatment by Medicare and beneficiaries.

**Figure 4.**
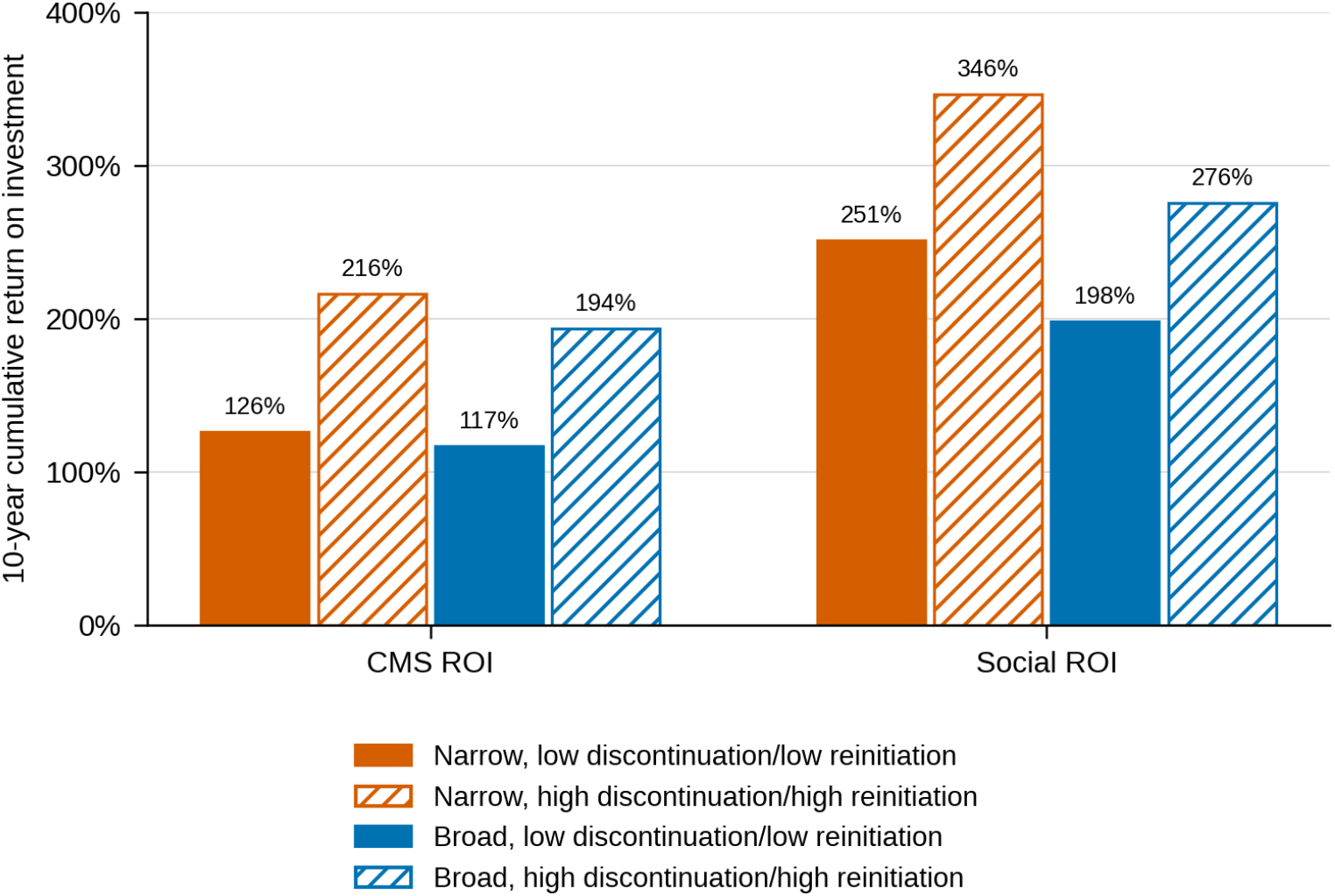
Ten-year cumulative ROI. **Notes:**Results are for Bridge-eligible beneficiaries and include diabetes drug price savings beginning in 2028.

The 10-year CMS ROI was positive in every scenario, ranging from 117% (low discontinuation/low reinitiation, broad) to 216% (high discontinuation/high reinitiation, narrow), and the social ROI ranged from 198% to 346%. Diabetes drug price savings are identical across scenarios and add to both returns without changing Bridge spending.

ROI was greater under high discontinuation/high reinitiation than under low discontinuation/low reinitiation, and greater under the narrow population definition than the broad definition. The first pattern reflects that when patients stay on treatment less, Bridge drug spending falls proportionally more than net social value, while the diabetes drug price savings are unaffected. However, the net social value remains highest under the low discontinuation and low reinitiation scenario. The second pattern reflects the healthier mix of initiators in the narrow population, who generate larger benefits at similar drug cost.

## Discussion

In our simulation of a Medicare GLP-1 Demonstration, expanded access generated positive cumulative net social value over 10 years under every combination of discontinuation, reinitiation, and eligibility assumptions we studied. Cumulative net social value by 2036 ranged from $44.3 billion to $58.5 billion, and Medicare’s net spending decreased in every scenario, with cumulative net savings of $21.2–27.1 billion by 2036. Whether the expanded coverage for GLP-1s pays for itself from Medicare’s perspective depends on the Bridge price extending to GLP-1s prescribed for diabetes and other conditions already covered in Part D. In our simulation, extending the lower price to these indications generated $31.9 billion in savings, producing a positive return for Medicare in every scenario (117–216%).

Our modeling and assumptions imply that persistence has less of an effect on value than cost. Under low discontinuation and low reinitiation, net social value was 2% higher in the broad population and 7% higher in the narrow population, while Medicare drug spending was roughly 40–50% higher. As a result, the ROIs were higher under high discontinuation and high reinitiation. This does not mean that discontinuation is beneficial; it means that, in our model, patients who stop treatment also stop generating spending, and the value forgone is smaller than the spending avoided. Because we paired each discontinuation scenario with the reinitiation rate observed in the corresponding real-world population, our results reflect the combined effect of stopping and restarting treatment, and we cannot separate the two. In real world use, treatment cycling or yoyoing may have detrimental health impacts that our model is not accounting for.^33,34^

Our results also suggest that the beneficiaries who initiate treatment mattered more than our assumptions around adherence. Net social value was 23–29% higher under the narrow definition, even though the number of initiators was identical. The broad population includes beneficiaries with prior myocardial infarction or stroke, who have already developed conditions that GLP-1s might prevent and face higher mortality, so each initiator gains less. This suggests that coverage criteria, in addition to price and persistence, can impact how much value a program delivers per dollar spent. This is consistent with our prior work showing that GLP-1s deliver the most lifetime value when started early, before chronic conditions develop^35^, so limiting access to sicker patients does not raise the program’s return on investment.

To put these results in context, CMS Administrator Mehmet Oz reported that approximately 600,000 beneficiaries had enrolled in Bridge by August 31, 2026.^25^ The Bridge Demonstration is scheduled to run for 1.5 years, from July 2026 through December 2027.^15^ Applying our simulated per-person results at 1.5 years to these early enrollees implies a net social value of $304-5165 per person, or $182-3,099 million in total depending on the scenario, and 21-30 thousand fewer years lived with diabetes. However, these results only capture a small share of the long-term value. Net social value was close to zero in the first year of our simulation and grew in every year thereafter (Figure 2), so an evaluation at 18 months versus 10 years of the demonstration would see mostly costs and little of the benefit that accrues afterward. CMS has said it will review program data before deciding whether to continue the expanded GLP-1 access offered through the Bridge demonstration^36^, but our results highlight the fact that data collected over an 18-month window will understate the benefits of GLP-1 treatment, because the health gains that drive value, including fewer years with diabetes, accumulate over time. Evaluations of the demonstration should therefore combine early utilization and persistence data with models that project longer-term outcomes.

Our study has several important limitations. First, simulated results depend on assumptions about uptake, persistence, and weight loss that remain uncertain. We use discontinuation rates from the published literature at the high and low ends of reported estimates, so that our scenarios bracket the range of plausible real-world persistence. Neither source was designed to describe Medicare beneficiaries with consistent low-cost access to GLP-1s. Second, the diabetes price savings assume that GLP-1 use among beneficiaries with diabetes grows only from 15% to 25% over the horizon, and that the price reductions extend to existing prescriptions starting in 2028. If use grows faster in this population or among other covered indications, savings from the lower price could be substantially larger.

Our model treats discontinuation as random with respect to treatment response, which likely understates the program’s value. In practice, the effect of GLP-1s on weight and related outcomes varies across patients, and those who discontinue are probably disproportionately those who benefit least. When these patients stop treatment, the program avoids paying for a drug that is doing little for them, and the average benefit among those who stay on treatment rises. Our discontinuation assumptions are therefore likely conservative, particularly in the high discontinuation scenarios, and the finding that persistence matters less for net social value than for cost would likely be stronger if discontinuation were modeled as selective. Future work should model heterogeneity in treatment response and the relationship between response and persistence.

In summary, expanded Medicare access to GLP-1s is projected to generate substantial social value across the range of discontinuation, reinitiation, and eligibility scenarios we modeled. Moreover, extending the lower demonstration pricing to GLP-1s prescribed for indications already covered in Part D would allow the demonstration to pay for itself from Medicare’s perspective.

## Data Availability

All data produced in the present study are available upon reasonable request to the authors.

https://schaeffer.usc.edu/wp-content/uploads/2025/02/FEM-Technical-Document.pdf

## Appendix

**Table A1.** Discontinuation and reinitiation parameters implemented in simulation.

|  | Rodriguez et al 2025 | UAS |
| --- | --- | --- |
| **Discontinuation probabilities (quarters since treatment initiation)** |  |  |
| First episode, quarters 1-4 | 14.5% | 5.7% |
| First episode, quarters 5-8 | 9.5% | 4.0% |
| First episode, quarters 9+ | 2.5% | 1.1% |
| Reinitiated episode | 2.5% | 1.1% |
| **Reinitiation probabilities (quarters since discontinuation)** |  |  |
| Quarters 1-7 | 57.3% | 30.0% |
| Quarters 8-15 | 10.4% | 6.0% |
| Quarters 16+ | 1.70% | 3.0% |

**Notes:** The table shows the probabilities used in the simulation to stop treatment (discontinuation) and to restart it after stopping (reinitiation). Discontinuation probabilities are quarterly. Reinitiation probabilities are two-year probabilities, assessed once every two years.
a. Discontinuation: Cumulative probabilities from Rodriguez et al 2025 are converted to annual probabilities, which are then converted to quarterly probabilities (we assume a constant hazard within each year). The probability depends on time since treatment initiation. We extrapolated the year 3+ probability using the rate of change between year 1 and year 2. The year 3+ probability is held constant for every later quarter. Discontinuation probabilities for subsequent treatment episodes are assumed to be the same as the year 3+ probabilities.
b. Reinitiation: A two-year reinitiation probability is derived from the UAS data (proxy is percentage of current users who have ever had a treatment gap of 3+ months). People who have stopped treatment are assessed for reinitiation once, at the start of each cycle. The probability depends on time since discontinuation at the point of assessment. A rate of change similar to Rodriguez et al 2025's is applied to the UAS probability to obtain year 3+ probabilities.
c. Repeat episodes: We limit the number of treatment episodes to two in a person's lifetime.

## Footnotes

a Beneficiaries with metabolic dysfunction-associated steatohepatitis (MASH) are eligible for Part D coverage, but we currently do not model MASH.

b Chronic kidney disease, prediabetes, and symptomatic peripheral artery disease are not incorporated in the current modeling approach.

c The Bridge demonstration began on July 1, 2026, so our simulation, which starts in 2027, does not capture the program’s first six months. Beneficiaries who began treatment in 2026 are instead treated as first-year initiators in 2027, so our estimates exclude costs and benefits accrued in 2026.

